# Feature pre-selection for the development of epigenetic biomarkers

**DOI:** 10.1101/2024.02.14.24302694

**Authors:** Yipeng Cheng, Christian Gieger, Archie Campbell, Andrew M McIntosh, Melanie Waldenberger, Daniel L McCartney, Riccardo E Marioni, Catalina A Vallejos

## Abstract

Over the last decade, a plethora of blood-based DNA methylation biomarkers have been developed to track differences in ageing, lifestyle, health, and biological outcomes. Typically, penalised regression models are used to generate these predictors, with hundreds or thousands of CpGs included as potential features. However, in such ultra high-dimensional settings, the effectiveness of these methods may be reduced.

Here, we introduce Related Trait-based Feature Screening (RTFS), a method for performing CpG pre-selection for incident disease prediction models by utilising associations between CpGs and health-related continuous traits. In a comparison with commonly used CpG pre-selection methods, we evaluate resulting downstream Cox proportional-hazards prediction models for 10-year type 2 diabetes (T2D) onset risk in Generation Scotland (n=18,414). The top performing models utilised incident T2D EWAS (AUC=0.881, PRAUC=0.279) and RTFS (AUC=0.877, PRAUC=0.277). The resulting models also improve prediction over a model using standard risk factors only (AUC=0.841, PRAUC=0.194) and replication was observed in the German-based KORA study (n=4,261)

RTFS is a flexible and generalisable framework that can help to refine biomarker development for incident disease outcomes.

## Introduction

Numerous studies have shown that levels of DNA methylation (DNAm) at various CpG sites can correlate with health-related traits, such as body mass index (BMI), smoking status [1], and incident diseases [2, 3, 4]. DNAm is an epigenetic modification whereby methyl groups are dynamically attached and removed at various genomic positions (often on the cytosine of a C-G dinucleotide; CpG) throughout an individual’s lifetime. Blood-based DNAm is of particular interest within cohort studies as its relatively non-intrusive sample procedure makes it potentially suitable for clinical biomarker development, enabling the development of risk prediction models (e.g. to predict incident disease).

A major challenge in developing these prediction models is the selection of relevant CpG sites for use as inputs. DNAm is commonly (and affordably) ascertained through the use of arrays including the Illumina Infinium HumanMethylation450 and EPIC arrays, which capture methylation information for *∼*450,000 and *∼*800,000 CpG sites, respectively [5, 6]. In contrast, cohort sizes tend to be limited to a few thousand individuals. This leads to an ultra high-dimensional setting in which the number of features or predictors (*p*) is much larger than the number of observations (*n*).

A typical approach to utilising high-dimensional data involves the application of penalised regression models, both for feature selection and prediction (see e.g. [1, 7, 8]). However, in ultra high-dimensional settings, the effectiveness of penalised regression may be reduced [9, 10, 11]. A two-stage process has previously been suggested to address this, where a pre-selection (screening) step is first applied to the data, before fitting penalised regression models [12, 10]. The purpose of the pre-selection is to broadly filter out irrelevant features to reduce the number of potential predictors to a size suitable for penalised regression (typically of polynomial order with respect to the sample size [11]).

One commonly used method for CpG pre-selection is variance-based filtering, whereby the top *k* CpGs are retained after ranking them by decreasing variance, where *k* is arbitrarily chosen. This method helps to remove invariant CpG sites, but its performance may be problematic, particularly with small effect and sample sizes [13]. Other approaches, based on the correlation of each feature with the outcome, have been proposed for continuous (e.g. [12]) and time-to-event data (e.g. [14]), but some of these may introduce problems related to post-selection inference [15] if the same data is used for screening and model fitting. An alternative is to use domain knowledge (e.g. from external data) to inform the screening. One such method involves pre-selecting CpGs that have previously shown associations with the outcome in Epigenome-Wide Association Studies (EWAS). If the associations have been found in an independent dataset, the chance of noise from spurious correlations with the outcome is reduced. However, the pre-selection is limited to marginal associations between each CpG site and the outcome and availability of EWAS results varies depending on the outcome. Another strategy, that can bypass the need for feature pre-selection, is the application of principal components analysis (PCA; or other dimensionality reduction techniques) to obtain a low-dimensional set of features (e.g. the first 100 principal components) to be used as inputs. The latter has been shown to potentially improve out-of-sample prediction in CpG-based models [16].

Here, we propose a Related Trait-based Feature Screening (RTFS) pipeline, using information about continuous traits that are related to the outcome of interest to perform feature pre-selection. For example, to predict time-to disease incidence, we selected a range of measurements (e.g. BMI, smoking, alcohol consumption) typically related to a broad set of health outcomes. Feature pre-selection can be then performed by applying e.g. penalised regression on the continuous traits, with lower sample size requirements than time-to-event data [17, 18, 19]. Power calculations for time-to-event data typically depend on the number of case events per feature, which is often small compared to the overall sample size. This is in contrast to the corresponding calculations for continuous traits which are based on the total number of data points per feature.

We apply RTFS and other popular CpG pre-selection methods to the Generation Scotland (GS) [20] cohort (*n* = 18, 414), one of the world’s largest studies including genome-wide DNAm data paired with linkage to electronic health records (EHR). We compare the performance of the different pre-selection methods as well as dimensionality reduction using PCA in the development of epigenetic scores (EpiScores) - weighted sums of CpG methylation values - used to predict time to incident type 2 diabetes (T2D). We show that RTFS is competitive with the top existing EWAS-based filtering approach, leading to an increase in predictive performance above standard T2D risk factors. We also show the predictive performance increases of the EpiScores compared to genetic risk factors using a T2D polygenic risk score (PRS). Finally, we validated the performance of resulting EpiScores derived from RTFS and incident T2D EWAS-based filtering in the KORA S4 cohort [21]. All analyses and results are reported in line with the TRIPOD checklist [22] for reproducibility purposes and can be found in **Supplementary File 1**. Analysis scripts are provided on GitHub at https://github.com/marioni-group/rtfs.

## Results

### RTFS

The proposed RTFS pipeline aims to aid feature pre-selection in ultra high-dimensional settings when developing prediction models for incident disease risk. We focus on time-to-event outcomes, but a similar pipeline could be applied to other types of outcomes (e.g. binary or counts). RTFS borrows information from a set of traits that are related to the outcome — or a broad set of outcomes — of interest. The process is illustrated in **Figure 1**. First, (linear) lasso regression models are trained with each of the (continuous) traits as the outcome. The resulting RTFS pre-selected CpG set consists of the union of CpG sites retained from any of the continuous trait models. Here, nineteen continuous traits were included in the RTFS pipeline: age, glucose, total cholesterol, high-density lipoprotein (HDL) cholesterol, sodium, potassium, urea, creatinine, BMI, waist-hip ratio, body fat percentage, systolic blood pressure, diastolic blood pressure, heart rate, forced expiratory volume (FEV), forced vital capacity (FVC), alcohol consumption, smoking and general cognitive ability. All of these were recorded at baseline (see **Methods**). For all the continuous traits, the in-sample predictive performance for the corresponding lasso model in the test set is given in **Supplementary Table 1**.

**Figure 1:**
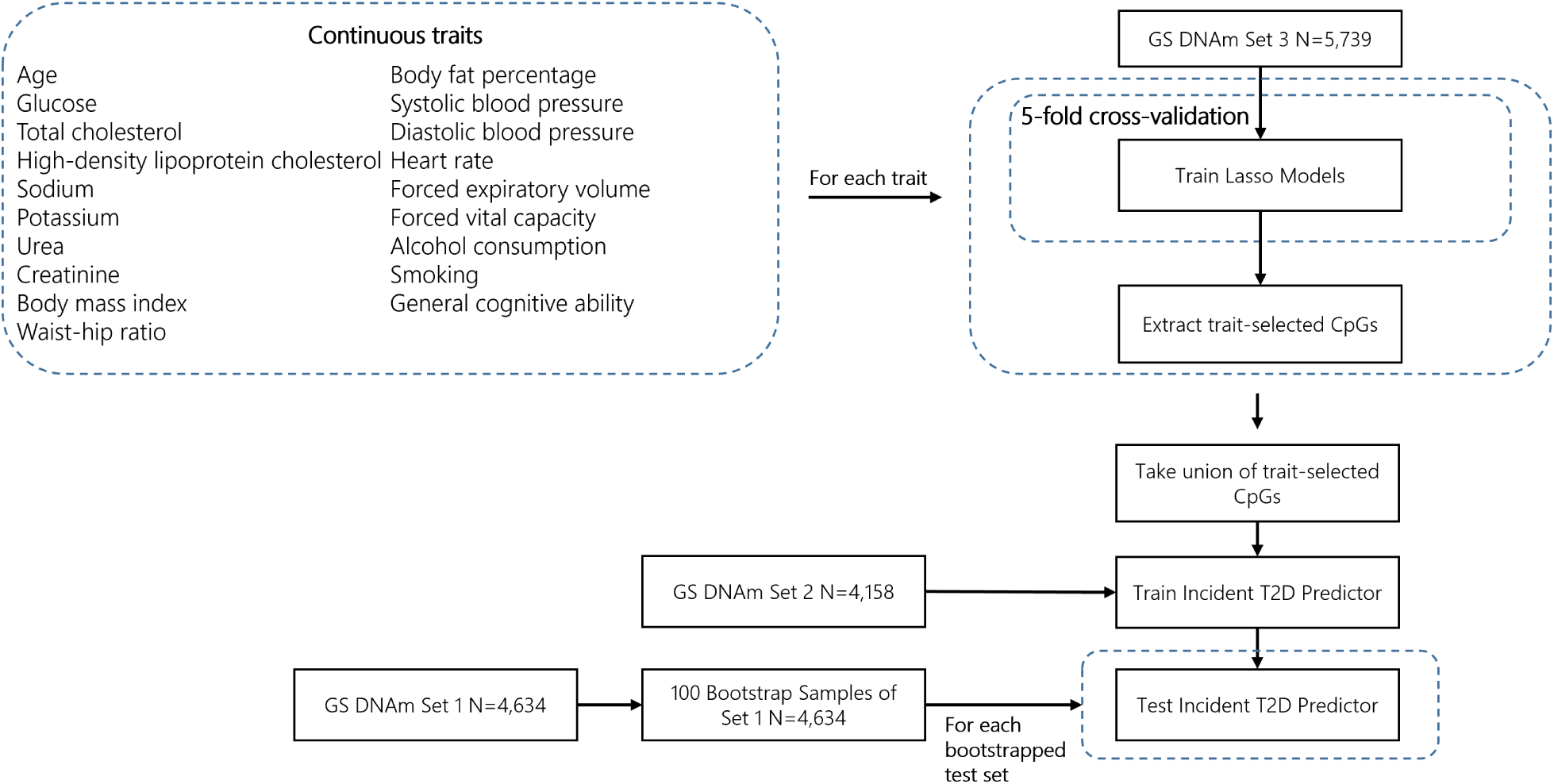
The RTFS pipeline applied to Generation Scotland.

### Cohort summary

After exclusions, our data consisted of 14,531 individuals from the GS cohort (see **Methods** and **Supplementary Figure 1**). This was divided into three non-overlapping sets: to train the trait-specific models (feature pre-selection set for RTFS only; *n* = 5, 739) as well as to train (*n* = 4, 158) and test (*n* = 4, 634) the incident T2D prediction model. After removal of missing values in the continuous traits, the pre-selection set consisted of between *n* = 4, 872 and *n* = 5, 739 individuals depending on the trait (see **Supplementary Table 2**). Summary information for the T2D training and test sets is shown in **Table 1** and **Supplementary Figure 2**. Both sets had a highly imbalanced case/control distribution with 3.2% (130/4,028) and 4.6% (213/4,634) having an incident T2D diagnosis in the training and test sets, respectively.

**Table 1:**
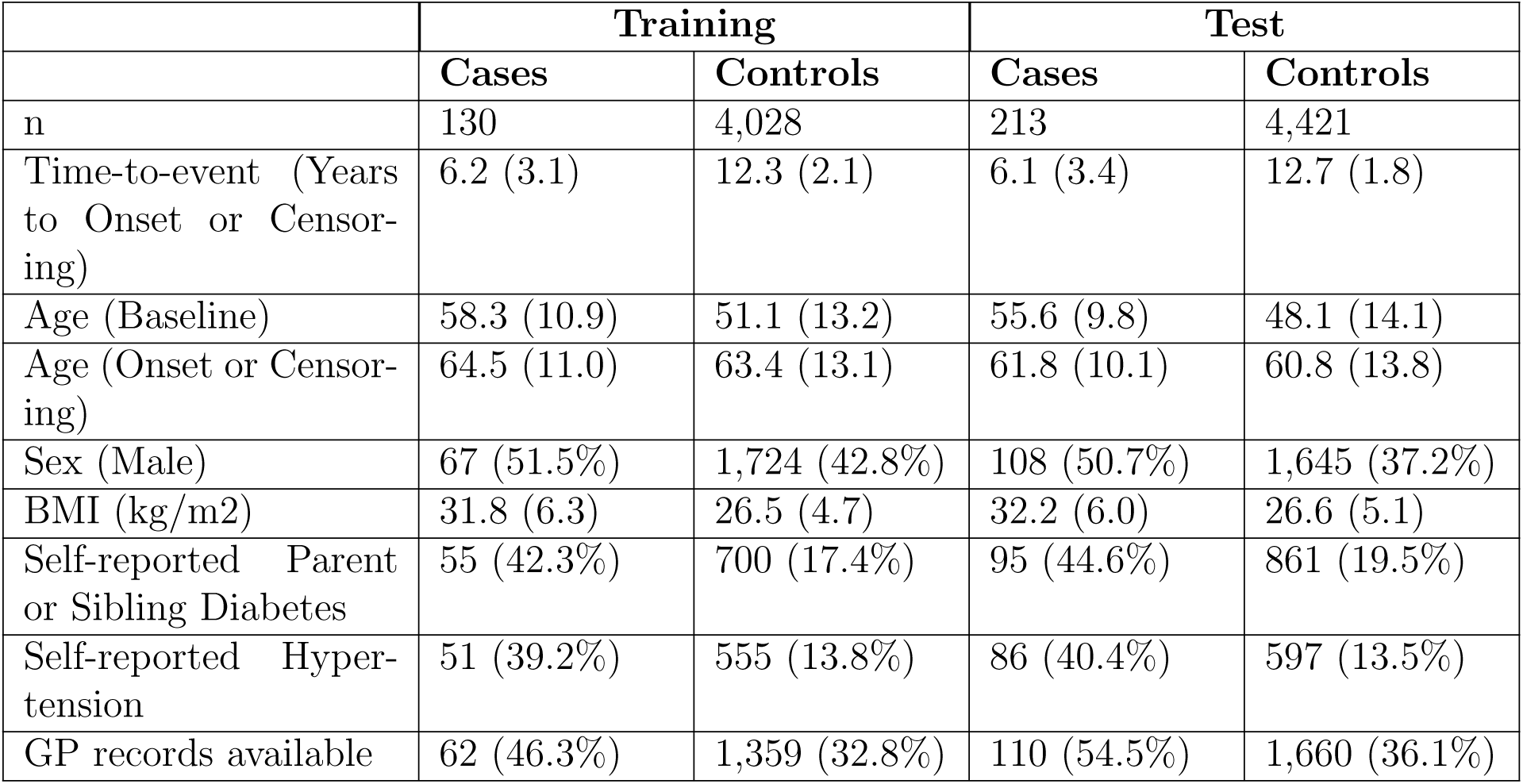
Summary details for the incident T2D training and test sets.

|  | Training |  | Test |  |
| --- | --- | --- | --- | --- |
|  | Cases | Controls | Cases | Controls |
| n | 130 | 4,028 | 213 | 4,421 |
| Time-to-event (Years to Onset or Censoring) | 6.2 (3.1) | 12.3 (2.1) | 6.1 (3.4) | 12.7 (1.8) |
| Age (Baseline) | 58.3 (10.9) | 51.1 (13.2) | 55.6 (9.8) | 48.1 (14.1) |
| Age (Onset or Censoring) | 64.5 (11.0) | 63.4 (13.1) | 61.8 (10.1) | 60.8 (13.8) |
| Sex (Male) | 67 (51.5%) | 1,724 (42.8%) | 108 (50.7%) | 1,645 (37.2%) |
| BMI (kg/m<sup>2</sup>) | 31.8 (6.3) | 26.5 (4.7) | 32.2 (6.0) | 26.6 (5.1) |
| Self-reported Parent or Sibling Diabetes | 55 (42.3%) | 700 (17.4%) | 95 (44.6%) | 861 (19.5%) |
| Self-reported Hypertension | 51 (39.2%) | 555 (13.8%) | 86 (40.4%) | 597 (13.5%) |
| GP records available | 62 (46.3%) | 1,359 (32.8%) | 110 (54.5%) | 1,660 (36.1%) |

### Prediction performance assessment

When assessing predictive performance in the test set, two types of outcomes were considered: prediction of time to incident T2D diagnosis and a binary outcome given by whether incident T2D occurred prior to 10 years after baseline (**Methods**). Predictive performance using the time to incident T2D diagnosis was assessed using C-index and Brier scores. C-index measures discrimination (agreement in the ranking between predicted risks and observed time-to-event values across pairs of individuals) while Brier scores give a measurement of both model calibration and discrimination at a given time point. In our experiments, Brier scores were evaluated at all integer time points from *t* = 1 to *t* = 10 years (inclusive). Binary outcome prediction performance was assessed using measures of discrimination - area under the receiver operating characteristics curve (AUC) and area under the precision recall curve (PRAUC). Calibration of the predictions generated by each model was also evaluated. Other measures, such as specificity and sensitivity, across a range of probability classification thresholds are also provided.

### Prediction of incident T2D using risk factors only

A Cox proportional-hazards (Cox PH) model in the test set using established risk factors (age, sex, BMI, hypertension and parent/sibling history of diabetes) as covariates (referred to as the risk factors-only model) had a C-index of 0.828 for time-to-event outcomes (Brier scores are shown in **Supplementary Table 3**). AUC and PRAUC were 0.841 and 0.194, respectively, when predicting if an incident T2D diagnosis occurred prior to 10 years after baseline.

### Prediction of incident T2D using risk factors and DNAm

We considered four methods for feature pre-selection (**Figure 2**; details in **Methods**): filtering to sites on the 450k array (henceforth referred to as the EPIC-450k intersection); filtering to the top 100k and 200k most variable CpGs; filtering to epigenome-wide significant CpGs from the EWAS literature (72 and 55 CpGs for incident and prevalent T2D, respectively); and filtering to the 5,468 RTFS CpGs identified from the lasso models on the continuous traits. We also considered applying PCA to the EPIC-450k intersection and to the top 200k most variable CpGs, with the PCs explaining a cumulative variance *>* 95% taken forward as features. This led to selecting 3,734 and 3,652 PCs, respectively. The greatest C-index values were achieved from using incident T2D EWAS-based filtering and RTFS (both 0.866). All C-index and Brier score values are shown in **Supplementary Table 3**. Incident T2D EWAS-based filtering and RTFS resulted in the lowest two Brier scores for all time points, suggesting that those methods consistently performed in the top two models in terms of calibration and case/control discrimination. **Table 2** shows the AUC and PRAUC values obtained from incremental Cox PH models corresponding to the addition of an EpiScore, derived from each pre-selection method or PCA, to the risk factors-only model. Incident T2D EWAS-based filtering achieved the highest AUC (0.881) and PRAUC (0.279). Corresponding ROC curves for the incident T2D EWAS-based filtering, RTFS and the risk factors-only models are shown in **Figure 3**. We evaluated the robustness of this ranking by considering the number of times each method was ranked in the top *n* methods across 1,000 bootstrap runs is plotted in **Figure 4**. Incident T2D EWAS-based filtering had the highest frequency of first rankings across the bootstraps in both AUC and PRAUC. RTFS also performed consistently well with both methods ranking in the top three in the majority of bootstraps.

Differences in model calibration between the incident T2D EWAS EpiScore model, RTFS EpiScore model and risk factors-only model are shown in **Supplementary Figure 4**. The incident T2D EWAS EpiScore and RTFS EpiScore models show stronger calibration performance when compared to the risk factors-only model. All three models plotted show underestimation of risk below a predicted probability of around 0.5 and overestimation of risk otherwise.

**Figure 2:**
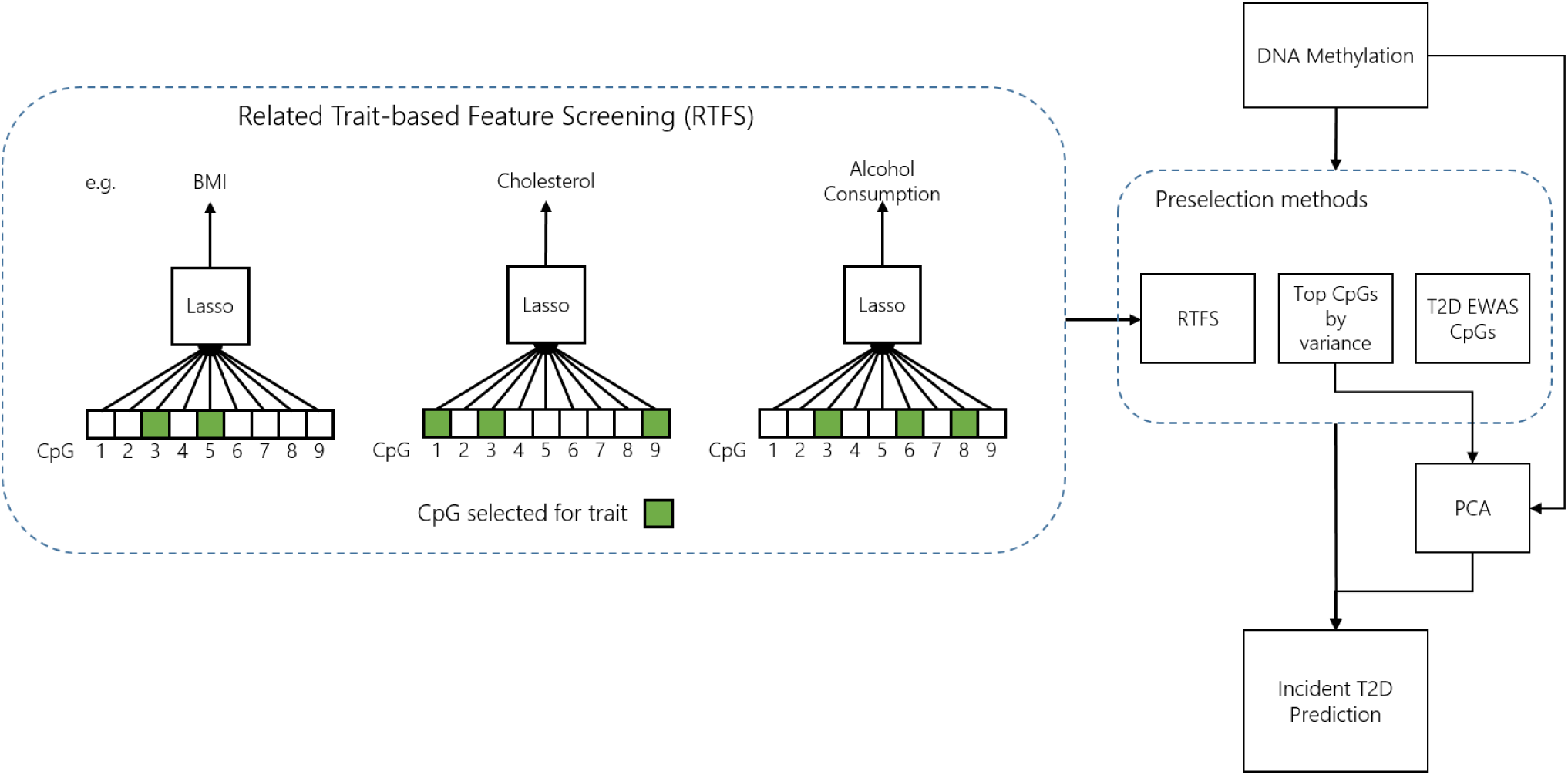
An overview of the pre-selection comparison pipeline.

**Figure 3:**
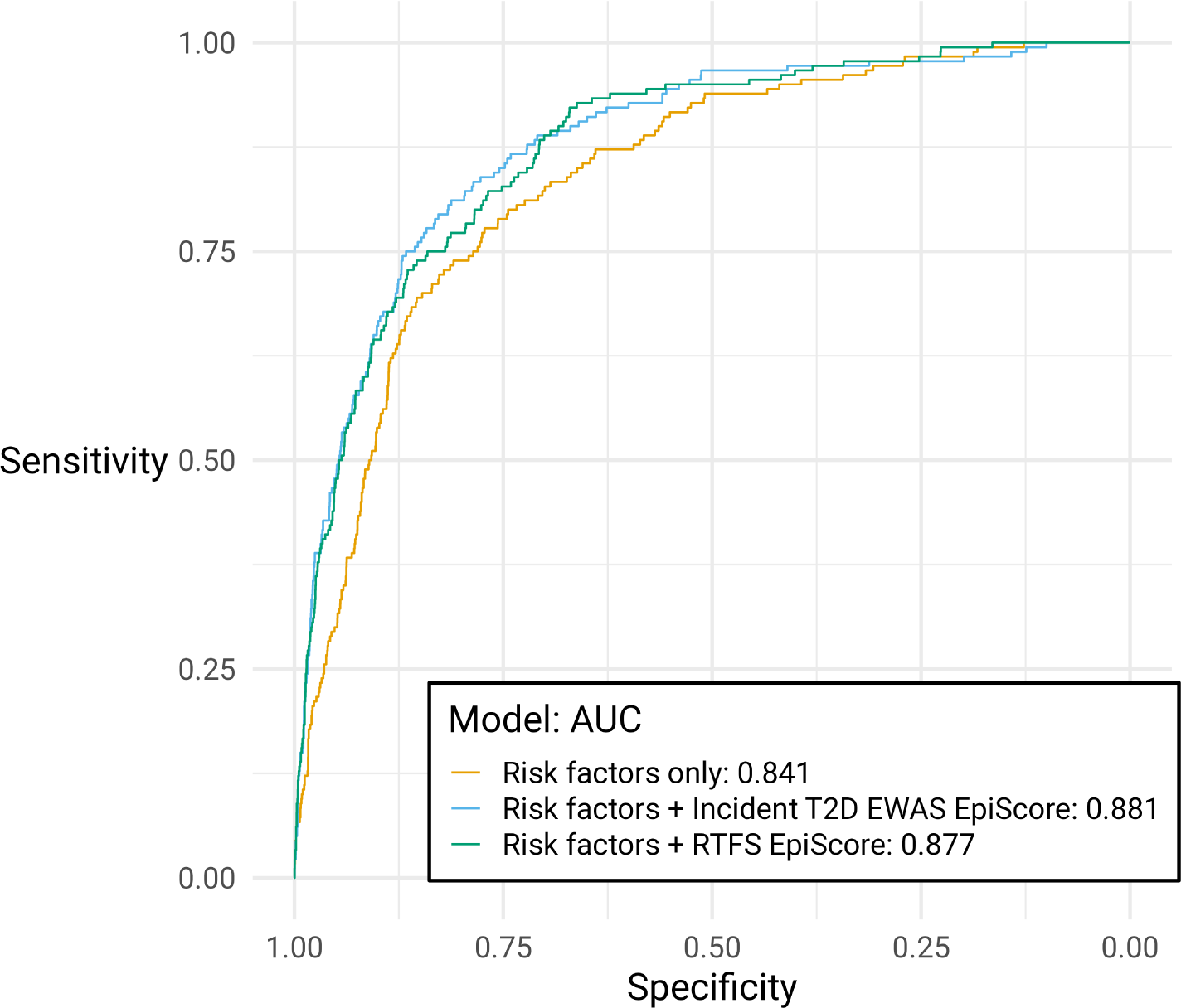
ROC curves for incremental incident T2D models. Results are shown for the model including risk factors only in addition to the models using RTFS and incident T2D EWAS-based filtering.

**Figure 4:**
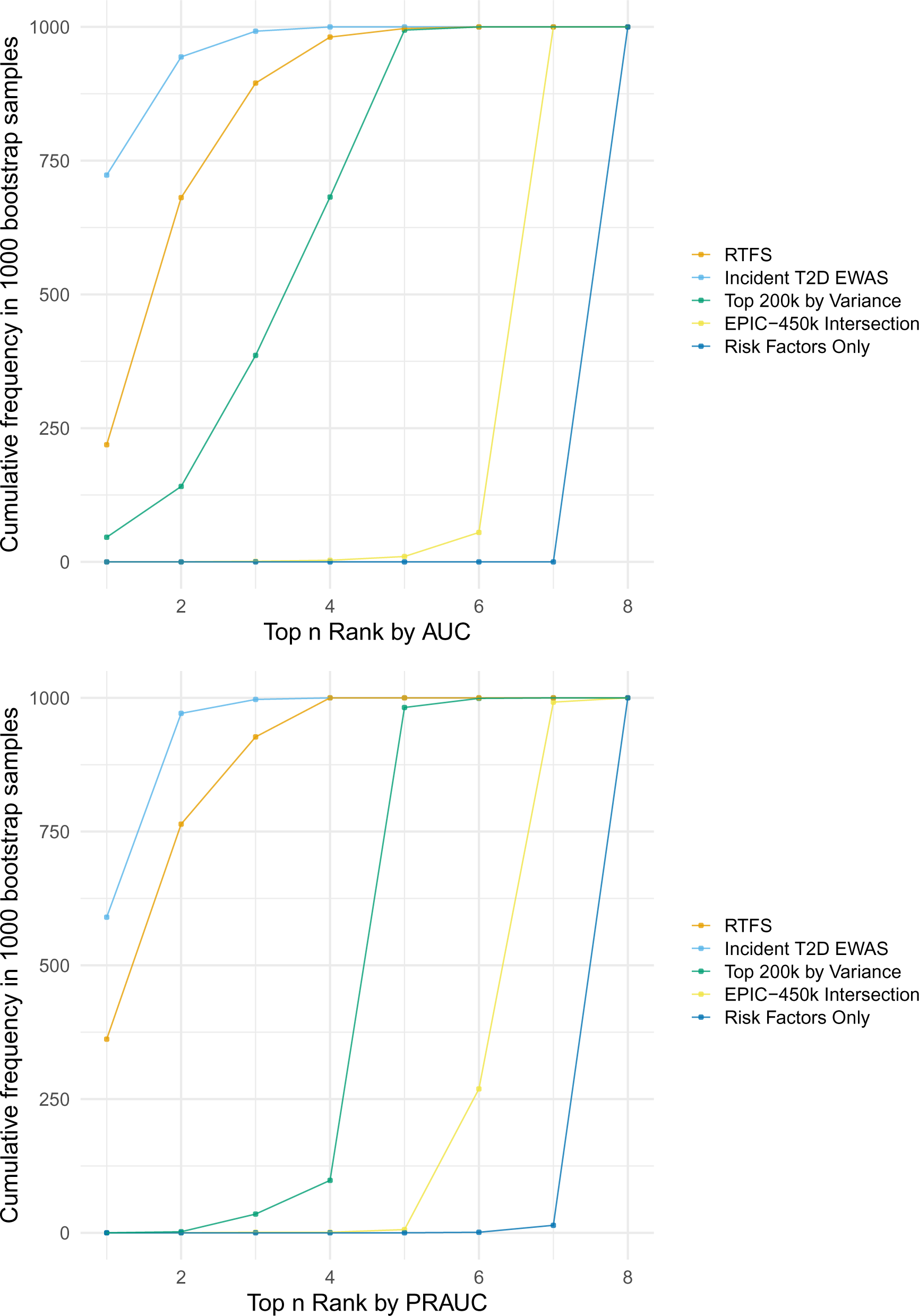
Rank order cumulative frequencies for AUC (top) and PRAUC (bottom) values for pre-selection methods across 1,000 bootstrap samples of the test set. The plots show, for each method, the number of bootstraps in which the method ranked in the top n in terms of their respective AUC/PRAUC. Models shown include: Related Trait-based Feature Selection (RTFS), Incident T2D EWAS, Top 200k by Variance, EPIC-450k Intersection, and Risk Factors Only.

**Table 2:** Incremental modelling performance metrics for each pre-selection / PCA result calculated in the GS test set.

| Incremental Model Variables (in addition to risk factors) | AUC | PRAUC | C-index | Alpha | Lambda |
| --- | --- | --- | --- | --- | --- |
| Risk Factors (RF) only | 0.841 | 0.194 | 0.828 | NA | NA |
| RF + PCA EPIC-450k EpiScore | 0.849 | 0.206 | 0.837 | 0.2 | 0.325 |
| RF + PCA Top 200k by Variance EpiScore | 0.853 | 0.205 | 0.841 | 0 | 9.716 |
| RF + EPIC-450k EpiScore | 0.855 | 0.208 | 0.841 | 0.8 | 0.014 |
| RF + PRS | 0.857 | 0.212 | 0.843 | NA | NA |
| RF + Top 100k by Variance EpiScore | 0.864 | 0.215 | 0.852 | 0.7 | 0.012 |
| RF + Prevalent T2D EWAS EpiScore | 0.869 | 0.255 | 0.858 | 0.6 | 0.004 |
| RF + Top 200k by Variance EpiScore | 0.872 | 0.233 | 0.860 | 0.5 | 0.017 |
| RF + Prevalent and Incident T2D EWAS EpiScore | 0.873 | 0.262 | 0.858 | 1 | 0.002 |
| RF + RTFS EpiScore | 0.877 | 0.277 | 0.866 | 0.5 | 0.012 |
| RF + Incident T2D EWAS EpiScore | 0.881 | 0.279 | 0.866 | 0.5 | 0.006 |
| RF + PRS + Incident T2D EWAS EpiScore | 0.892 | 0.302 | 0.876 | 0.5 | 0.006 |

**Supplementary Figure 3** shows how confusion matrix values vary across the probability classification threshold range for the risk factors-only, RTFS and the incident T2D EWAS-based filtering EpiScore model in the test set. Overall, the incident T2D EWAS-based filtering and RTFS EpiScore model improve the classification of cases with respect to the risk factors-only model (increase in true positives and decrease in false negatives) while showing a slight decrease in the correct classification of controls. The differences in correctly classified individuals in terms of sensitivity, specificity, positive predictive value (PPV) and negative predictive value (NPV) between the RTFS EpiScore and risk factors-only models are also given in **Supplementary Table 4**.

### Comparison of incident T2D EpiScore and polygenic risk score prediction performance

To assess the added value of the EpiScores against genetic risk factors on predictive performance, two additional Cox PH models were fit to the GS test set that included a polygenic risk score (PRS) for incident T2D [23]. These consisted of a model using the standard risk factors plus the PRS, as well as a second model which also included the EpiScore derived from incident T2D EWAS-based filtering (the top performing pre-selection method). These two models showed AUC values of 0.857 and 0.892 respectively. PRAUC values were 0.212 and 0.302 and C-index values were 0.843 and 0.876. The PRS gave a smaller increase in each of these metrics above standard risk factors compared to the incident T2D EWAS EpiScore (AUC=0.881, PRAUC=0.279, C-index=0.866); however, without pre-selection of CpG sites, the EpiScore gives smaller increases (EPIC-450k EpiScore AUC=0.855, PRAUC=0.208, C-index=0.841. The largest increase was given when using both the PRS and EpiScore in the model, showing additive increases from both scores over using risk factors only.

### Validation of RTFS and EPIC-450k intersection EpiScores in the KORA S4 cohort

Performance in KORA S4 was only evaluated for the binary T2D incidence outcome (diagnosis within 10 years of baseline date) as time to T2D diagnosis data was not avail-able. The logistic risk factors-only model fit to the KORA S4 cohort showed an AUC and PRAUC of 0.797 and 0.294, respectively. The logistic models including risk factors plus either the RTFS or EPIC-450k incident T2D EpiScore resulted in AUCs of 0.806 and 0.798, respectively. Corresponding PRAUC values were 0.295 and 0.293 (see **Supplementary Table 5**.

### Overlap of pre-selected CpG sites

The continuous trait lasso models in RTFS selected between 49 and 864 CpG sites per trait (5,468 in the union). **Figure 5** shows the number of CpG sites selected for each trait and the selection overlap between traits. This shows that the majority of CpG sites were selected exclusively for a single trait. Notable overlaps were present between BMI, waist-to-hip ratio and body fat as well as between systolic and diastolic blood pressure.

**Supplementary Figure 5** shows the number of CpGs pre-selected across all methods and their overlap. Over half of the RTFS pre-selected CpGs were not in the top 200,000 CpGs by variance. Additionally, a small proportion of the incident and prevalent EWAS CpGs overlapped with the RTFS CpGs (see **Supplementary Figure 6**)

**Figure 5:**
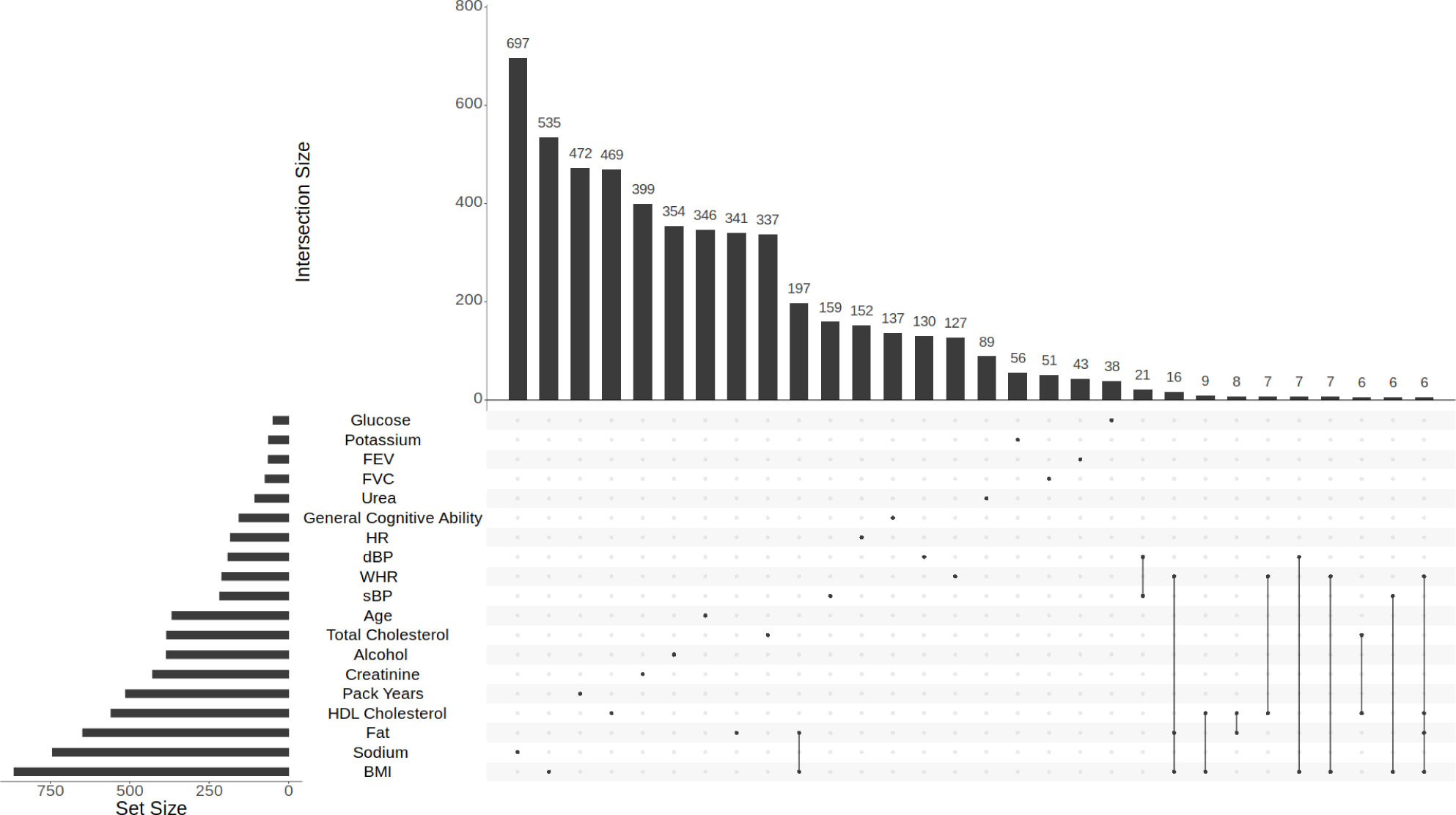
UpSet plot showing number of CpGs selected for each continuous trait and overlaps between traits. The frequency of the top 30 trait combinations are shown. Each column represents the number of CpGs pre-selected for the corresponding specific combination of traits. This was generated with the ”distinct” option, meaning the presence or absence of a point in a column explicitly corresponds to the presence or absence of the corresponding trait in the set

## Discussion

In this study, we explored the use of different feature pre-selection methods in the context of ultra-high dimensional DNAm data (where the number of features largely exceeds the number of observations). We introduce RTFS, which borrows information from a broad set of health-related traits to identify a suitable set of CpG sites that can be used as input in the development of risk prediction models for incident disease. Using type 2 diabetes as a case study, we compared the performance of RTFS against a range of other commonly-applied CpG pre-selection (and dimensionality reduction) approaches. Consistent with [24], the inclusion of an EpiScore generally improved discrimination performance with respect to the standard risk factors-only model. However, the improvement was not uniform across the different methods: with only marginal improvements in the absence of feature pre-selection when training the EpiScore. Our analysis also shows that EpiScores can improve predictive performance compared to the use of genetic information via an existing incident T2D PRS.

Incident T2D EWAS-based filtering resulted in the highest AUC (0.881) and PRAUC (0.279) for 10-year incident disease prediction, with a notable increase in the correct classification of cases and a small decrease in correct classification of controls.External validation in KORA supported this, although it showed smaller improvements compared to an earlier study which used a larger training set for the incident T2D EpiScores [24]. While filtering to significant CpG sites from an incident T2D EWAS study was the highest performing model, it is reliant on the existence of large-scale EWAS studies for T2D, something that may not be generally available for other diseases of interest. RTFS bypasses this requirement and led to similar performance metrics (AUC = 0877; PRAUC = 0.277). It was also consistently ranked amongst the top performing models in our bootstrap experiments. Additionally, the continuous traits used for RTFS were primarily general health-related measures and not necessarily specific to T2D. Therefore, the resulting set of RTFS CpGs may be applicable to other diseases and could potentially be used as a general panel of morbidity-related sites for risk prediction.

We used a pre-specified set of continuous trait to perform CpG pre-selection in RTFS. While we evaluated the predictive performance of each trait-specific lasso model, future studies could investigate the impact of including or excluding continuous traits e.g. based on a range of different performance thresholds. Additional studies could also investigate other variable selection methods for RTFS continuous traits, for example using elastic-net [25], as well as more general methods for risk prediction (e.g. random survival forests[26]). Future studies could also consider using DNAm-based predictions for each trait directly as predictors in downstream models, similar to previous approaches (e.g. the protein EpiScores in [27] or the approach used to develop the GrimAge epigenetic clock [28]).

Access to the GS cohort enabled us to demonstrate the use of RTFS in one of the largest cohorts of its kind — with three independently-processed sets of DNAm data, which allowed for separate training, testing and RTFS pre-selection datasets. In addition, comprehensive information on incident T2D diagnoses was available through extensive linkage to electronic health records. Availability of both genetic and epigenetic data allowed for a direct performance comparison between risk scores derived from each data source and showed the benefit of using DNAm data, which can better reflect health-associated changes within individuals’ lifetimes. While the inclusion of DNAm resulted in considerable predictive performance increases compared to using risk factors only in the GS test set, these differences were small when applied to the KORA validation cohort. The generalisability of our results is limited by the characteristics of the GS cohort: GS participants are generally healthier, wealthier and have a different age-sex distribution to the general population [29]. Similarities in these socio-demographic characteristics within GS may have resulted in positive bias in the performance of RTFS. Given that the models including DNAm data with and without CpG preselection both showed small performances differences when compared to a risk factors-only model in KORA, further work could explore the impact of factors such as the number of incident cases and availability of primary versus secondary care data for T2D disease ascertainment. Additionally, both the development and validation cohorts consisted of individuals from predominantly white European ancestries. Further validation is required to evaluate the generalisability of RTFS to other populations and genetic ancestries.

In conclusion, our study reiterated the need for pre-selection as an important step in DNAm-based risk prediction models. We introduced and evaluated an effective pre-selection method, RTFS, utilising information from health-related traits with the potential for application in predictive models for other incident diseases in future studies.

## Methods

### Generation Scotland (GS) DNAm data

The data used for this study were from the Generation Scotland (GS) cohort, recruited from across Scotland between 2006 and 2011. This consists of 23,960 volunteers aged 18-99 at baseline (recruitment date). Of these, 18,414 have genome-wide DNAm data available, ascertained from blood samples taken at baseline. DNAm quality control is detailed in [24]. DNAm measurements were obtained in three large sets, processed in 2017 (set 1, *n* = 5, 087), 2019 (set 2, *n* = 4, 450) and 2021 (set 3, *n* = 8, 877). Set 2 was used as the training set for incident T2D and set 3 was used for feature pre-selection. Set 1 was used as the test set for incident T2D. Sets 1 and 3 contained related individuals (genetic relationship matrix (GRM) threshold *>* 0.05), both within and between sets. There were also related individuals between sets 2 and 3. To avoid the presence of families with individuals across the training and test sets, individuals in set 3 with a family member present in set 1 were excluded from the analyses (*n_excluded_* = 3, 138). To maintain compatibility with previous studies using the Illumina 450K array, the CpGs were filtered to those present in both the 450K and EPIC arrays (EPIC-450k intersection).

A range of traits were also recorded at baseline via questionnaire or clinical appointment. These included (units listed within parenthesis): age (*years*), glucose (*millimoles per litre; mmol/L*), total cholesterol (*mmol/L*), high-density lipoprotein (HDL) cholesterol (*mmol/L*), sodium (*mmol/L*), potassium (*mmol/L*), urea (*mmol/L*), creatinine (*mmol/L*), BMI (*kg/m*^2^), waist-hip ratio, body fat percentage, systolic blood pressure (*millimetres of mercury; mmHg*), diastolic blood pressure (*mmHg*), heart rate (*beats per minute; bpm*), forced expiratory volume (FEV) (*L*), forced vital capacity (FVC) (*L*), alcohol consumption (*units/week*), smoking (*pack years*) and general cognitive ability. The latter was defined as the first unrotated principal component from a PCA of four cognitive tests (logical memory, digit symbol, verbal fluency and vocabulary), scaled to mean of 0 and standard deviation of 1 [30].

### Continuous trait preprocessing

The 19 baseline traits listed above were used as the continuous traits for RTFS. These were processed separately within each data set, to remove outliers, and to regress out age and sex (after trait-specific transformation, if applied). Trait-specific transformation included adding 1 to each value of alcohol consumption and smoking, prior to a natural log-transform. Glucose and BMI were also log-transformed. Outliers were defined as points greater than 4 standard deviations away from the mean. This is with the exception of BMI for which outliers were defined as a BMI *<* 18 or BMI *>* 50. A linear regression model with *age*, *age*^2^ (to include non-linear effects with age) and *sex* as covariates was then fit to each continuous trait. The resulting residuals were kept for further analyses. For FEV and FVC, height was also included in the linear regression. Missing values in each continuous trait were treated as missing-at-random and corresponding individuals were removed from the training set when fitting the predictive model for the respective trait. The number of missing values for each trait is given in **Supplementary Table 2.**

### Time to incident Type 2 Diabetes (T2D)

History of disease diagnoses (prevalent and incident) was ascertained via data linkage to NHS Scotland health records. Secondary care (hospital) records from January 1980 to April 2022 were available for all subjects, with disease diagnoses encoded using ICD-9/10. Due to restricted consent from data controllers, only partial linkage to primary care (general practice; GP) records was available (a subset of general practice centres were unable to provide data): only available for 35% (*n* = 3, 191, *n_training_* = 1, 421*, n_test_* = 1, 770) of individuals in the incident T2D training and test sets. Primary care records cover the period from January 1980 to October 2020 and use Read2 codes to record disease diagnoses.

Hospital record-derived prevalent and incident T2D cases were defined as individuals with an E11* ICD-10 code or 250.0/250.1 ICD-9 code. GP record-derived cases were defined using a set of diabetes-related Read2 codes. A full list of ICD-9/10 and Read2 codes is provided in **Supplementary Table 6**. Type 1 and juvenile diabetes cases were treated as controls (no T2D). Additional prevalent cases were identified from self-reported history in a baseline questionnaire. All prevalent cases were removed. For incident cases, time-to-event (*years*) was calculated as the time from baseline to disease onset (first T2D record) for cases, and to censoring for controls. Controls were censored at the latest date of available hospital records (April 2022) or time-to-death, whichever happened sooner.

For the individuals with both primary and secondary care records, a comparison between time-to-event outcomes derived from hospital and GP records was used to assess possible delays in hospital diagnoses. As a sensitivity analysis (RTFS only), the end of GP follow-up (October 2020) was also considered as a censoring date for those individuals (in the absence of a hospital diagnosis). **Supplementary Table 7** shows the AUCs, PRAUCs and incremental Cox PH model coefficient estimates from this analysis. Differences between the two outcome derivations were minor in terms of both the discrimination metrics and coefficient estimates.

### T2D risk factors

Risk factors used in the incremental EpiScore models included age, sex, BMI, hypertension and parent/sibling history of diabetes. Hypertension and parent/sibling history of diabetes were defined as self-reported in the baseline questionnaires. While many T2D risk factors have been identified, we based these on the most utilised factors in existing risk scores according to [31]. These five risk factors were used as variables in the risk factors-only model.

### Related Trait-based Feature Screening (RTFS)

Linear lasso [32] was applied to each continuous trait (after pre-processing) using set 3. Lasso is a penalised regression method which shrinks regression coefficients to be small, forcing some to be exactly equal to zero. As such, it performs feature selection by keeping only the features with non-zero coefficients. The strength of the penalty is controlled by a hyper-parameter *λ*. Five-fold cross-validation was used to select *λ*, to minimise the mean squared-error of out-of-sample predictions. Lasso models were fit using the glmnet R package version 4.1-1 [33]. For computational efficiency, the top 200,000 sites with the highest marginal variance were used as inputs. The union of lasso-selected CpGs from the final set of continuous trait models (hereafter referred to as the RTFS set) were used as input to predict time-to T2D incidence.

As RTFS performs feature pre-selection via linear models applied to a set of continuous traits, it implicitly assumes that each trait can be predicted by a linear combination of CpGs. The predictive ability of DNAm for each trait was quantified in a test set (GS set 1) using the percentage of variance explained *R*^2^.

### Alternative approaches for feature pre-selection

Initially, the EPIC-450k intersection set was used without pre-selection. Two commonly used approaches for feature pre-selection were then considered as an alternative to RTFS:

#### Highest variance

The per-feature variance is calculated, and the top *p* features with the highest variance in set 3 are pre-selected. For the T2D analysis, we used *p* = 100, 000 and *p* = 200, 000.

#### EWAS-based filtering

Existing EWAS analysis for incident or prevalent disease were used to pre-select CpGs. For the T2D analysis, CpGs identified to be statistically significant by two recent large meta analyses for incident and prevalent T2D were included. The first study [34] consisted of five European cohorts (*N_cases_* = 1, 250*, N_controls_* = 1, 950) and identified 76 differentially methylated CpG sites (*p <* 1.1*×*10*^−^*^7^) for incident T2D. After filtering these to those present in the EPIC-450k intersection, 72 CpG sites remained. The second [35] consisted of four European cohorts (*N_cases_* = 340*, N_controls_* = 3, 088) identifying 58 differentially methylated CpG sites (*p <* 1.0 *×* 10*^−^*^5^) for prevalent T2D (55 post EPIC-450k intersection filtering). The full list of CpG sites identified from the two studies is shown in **Supplementary Table 8**.

### Dimensionality reduction

As an alternative to feature pre-selection, we also explored whether dimensionality reduction techniques can be used to create a low-dimensional set of features to be used as input when predicting T2D. Here, we focus on PCA (as in [16]). In this study, we applied PCA (in set 2) to the 450k-EPIC intersection and the top 200k CpGs by variance. PCs were ordered by the variance explained in set 2 and the top PCs required to explain 95% of the variance were kept for the final T2D model.

### Incident T2D EpiScore

Using the CpGs identified by each pre-selection method (or top PCs, where appropriate), a Cox PH elastic-net model [36] was fit to the set 2 DNAm data (training set) using time-to-T2D incidence as the outcome. Similar to lasso, elastic-net provides a regularised model fit, reducing overfitting. The strength of the regularization is controlled by hyper-parameters *λ* and *α*. If *α >* 0, the model performs feature selection by setting a subset of coefficients to 0. Hyperparameters were selected using 9-fold cross-validation. Lambda was optimised using the cv.glmnet function. Alpha was selected by testing values between 0 and 1 (inclusive) in increments of 0.1 and selecting the value which maximised mean partial-likelihood across the nine folds. The linear predictor from the resulting Cox PH elastic-net model was defined as an incident T2D EpiScore.

### T2D Incremental Modelling

The incident T2D EpiScores obtained after applying each feature pre-selection (or dimensionality reduction) method were subsequently applied to set 1 (test set) in an incremental modelling approach. Firstly, a risk factors-only model was fit using a Cox PH model and a set of known T2D risk factors (listed above) as predictors. For each pre-selection method, Cox PH models were then fit using the same variables as the risk factors-only model plus the corresponding EpiScore. Full details on the incremental modelling calculations are given in the **Supplementary Note**.

### Incident T2D Polygenic Risk Score

To compare the differences in predictive performance of the EpiScores to genetic risk factors, two additional Cox PH models were fit in the test. The first included the standard risk factors plus a T2D PRS [23]. The second included these same variables plus the top performing incident T2D EpiScore (incident T2D EWAS-based filtering).

### Predictive performance evaluation

Predictive performance for each of the Cox PH models above was evaluated on the test set. Two types of prediction outcomes were used: time-to-T2D diagnosis and a binary outcome defined by whether a T2D diagnosis was recorded within 10 years from baseline. For the time-to-T2D outcome, C-index and Brier scores were calculated using the SurvMetrics R package (version 0.5.0) [37]. C-index gives a measure of discrimination for a model, defined as proportion of concordant pairs of individuals predicted by the model. This value is between 0 and 1 (inclusive) with higher scores representing better discrimination. A pair of individuals is concordant if the individual with the smaller time-to-event is given a greater risk by the model. The Brier score measures both discrimination and calibration, calculated as the mean square difference between the true classes (i.e. whether a T2D diagnosis has occurred) and the predicted probabilities at a given time point. Brier scores range between 0 and 1 (inclusive) and lower scores represent better discrimination and calibration. Brier scores were evaluated at each integer time point from *t* = 1 to *t* = 10.

For the binary 10-year T2D onset outcome, predictions were calculated as one minus the estimated 10-year survival probability. This calculation was based on the Breslow estimator [38] for the cumulative baseline hazard. This calculation is detailed in the **Supplementary Note**. Censored individuals were defined as controls when assessing predictive performance. Discrimination metrics including area under the receiver operating characteristics curve (AUC) and the area under the precision-recall curve (PRAUC) were compared.

Confusion matrix metrics were also assessed by calculating the number of true/false positives/negatives using the ten-year onset probabilities and a range of discrimination thresholds between 0 and 1, in increments of 0.1. Calibration was assessed by plotting loess calibration curves using the valProbggplot function in the CalibrationCurves R package (version 2.0.0) [39]. These show the observed event proportions plotted against the predicted event probabilities.

To assess the robustness of the relative rankings for the pre-selection methods, the incremental modelling was repeated in 1,000 bootstrap samples of the test set for each EpiScore model. For each bootstrap sample, the EpiScore and risk factors-only models (**Table 2**) were ranked based on their AUC and PRAUC estimates. The number of times that each method was included in the top *n* ranks was calculated (*n* = 1*, . . .,* 10).

### Overlap of pre-selected CpG sites

The sets of CpGs selected across the continuous trait lasso models were analysed using an UpSet plot (UpSetR R package, version 1.4.0 [40]) showing, the number of CpG sites selected across all of the traits in each combination of continuous traits. The same visualisation method was used for analysing the overlap between CpGs selected using each pre-selection method.

### Validation of RTFS and EPIC-450k intersection EpiScores in the KORA S4 cohort

The incident T2D EpiScores derived from the RTFS and EPIC-450k intersection CpGs were validated in a subset of the German-based KORA S4 cohort, which consisted of 1,451 individuals aged 25-74 years and recruited in southern Germany. The subset was defined by individuals with DNAm and incident T2D data available, after removing prevalent cases at baseline. Missing CpG values in the DNAm data were mean-imputed and individuals with missing health measures were removed from the dataset.

The prediction outcome was defined as the occurrence of a T2D diagnosis within 10 years after individuals’ baseline date. A time-to-event outcome was not used for validation as time-to-T2D diagnosis data was not available in KORA S4.

Validation was performed using incremental logistic models. Firstly, a risk factors-only model was fit to the KORA S4 subset using age, sex, BMI, hypertension and parent history of diabetes as variables. Then, two additional logistic models were fit using the risk factors plus each of the RTFS and EPIC-450k intersection EpiScores. Prediction performance was evaluated by calculating AUC and PRAUC for each of the three models.

The incident T2D EWAS EpiScore was not validated in KORA S4 as the corresponding EWAS meta-analysis included KORA participants.

Additional details of participant follow-up, ascertainment of incident T2D diagnoses and preprocessing numbers are provided in the **Supplementary Note**.

## Data Availability

According to the terms of consent for Generation Scotland participants, access to data must be reviewed by the Generation Scotland Access Committee. Applications should be made to. The informed consent given by the KORA S4 study participants does not cover data posting in public databases. However, data are available upon request from the KORA Project Application Self-Service Tool. Data requests can be submitted online (https://epi.helmholtz-muenchen.de/) and are subject to approval by the KORA board.

## Code and data sharing

Analysis scripts for this study are available at https://github.com/marioni-group/rtfs. According to the terms of consent for Generation Scotland participants, access to data must be reviewed by the Generation Scotland Access Committee. Applications should be made to. The informed consent given by the KORA S4 study participants does not cover data posting in public databases. However, data are available upon request from the KORA Project Application Self-Service Tool. Data requests can be submitted online (https://epi.helmholtz-muenchen.de/) and are subject to approval by the KORA board.

## Ethics

All components of Generation Scotland received ethical approval from the NHS Tayside Committee on Medical Research Ethics (REC Reference Number: 05/S1401/89). Generation Scotland has also been granted Research Tissue Bank status by the East of Scotland Research Ethics Service (REC Reference Number: 20-ES-0021), providing generic ethical approval for a wide range of uses within medical research. Written, informed consent was provided by Generation Scotland participants.

The KORA studies were approved by the Ethics Committee of the Bavarian Medical Association (Bayerische Landesärztekammer; S4: #99186) and were conducted according to the principles expressed in the Declaration of Helsinki. All study participants gave their written informed consent.

## Acknowledgements

This research was funded in whole, or in part, by the Wellcome Trust [104036/Z/14/Z, 108890/Z/15/Z, 216767/Z/19/Z]. For the purpose of open access, the author has applied a CC BY public copyright licence to any Author Accepted Manuscript version arising from this submission. Generation Scotland received core support from the Chief Scientist Office of the Scottish Government Health Directorates (CZD/16/6) and the Scottish Funding Council (HR03006) and is currently supported by the Wellcome Trust (216767/Z/19/Z). DNA methylation profiling of the Generation Scotland samples was carried out by the Genetics Core Laboratory at the Edinburgh Clinical Research Facility, Edinburgh, Scotland and was funded by the Medical Research Council UK and the Wellcome Trust (Wellcome Trust Strategic Award ”STratifying Resilience and Depression Longitudinally” (STRADL; Reference 104036/Z/14/Z). The DNA methylation data assayed for Generation Scotland was partially funded by a 2018 NARSAD Young Investigator Grant from the Brain & Behavior Research Foundation (Ref: 27404; awardee: Dr David M Howard) and by a JMAS SIM fellowship from the Royal College of Physicians of Edinburgh (Awardee: Dr Heather C Whalley). Y.C. is supported by the University of Edinburgh and University of Helsinki joint PhD program in Human Genomics. C.A.V. is a Chancellor’s Fellow funded by the University of Edinburgh. R.E.M. is supported by Alzheimer’s Society major project grant AS-PG-19b-010.

The KORA study was initiated and financed by the Helmholtz Zentrum München – German Research Center for Environmental Health, which is funded by the German Federal Ministry of Education and Research (BMBF) and by the State of Bavaria. Furthermore, KORA research has been supported within the Munich Center of Health Sciences (MC-Health), Ludwig-Maximilians-Universität, as part of LMUinnovativ and is supported by the DZHK (German Centre for Cardiovascular Research). The KORA study is funded by the Bavarian State Ministry of Health and Care through the research project DigiMed Bayern (www.digimed-bayern.de).

## Supplementary Materials

**Supplementary Figure 1:**
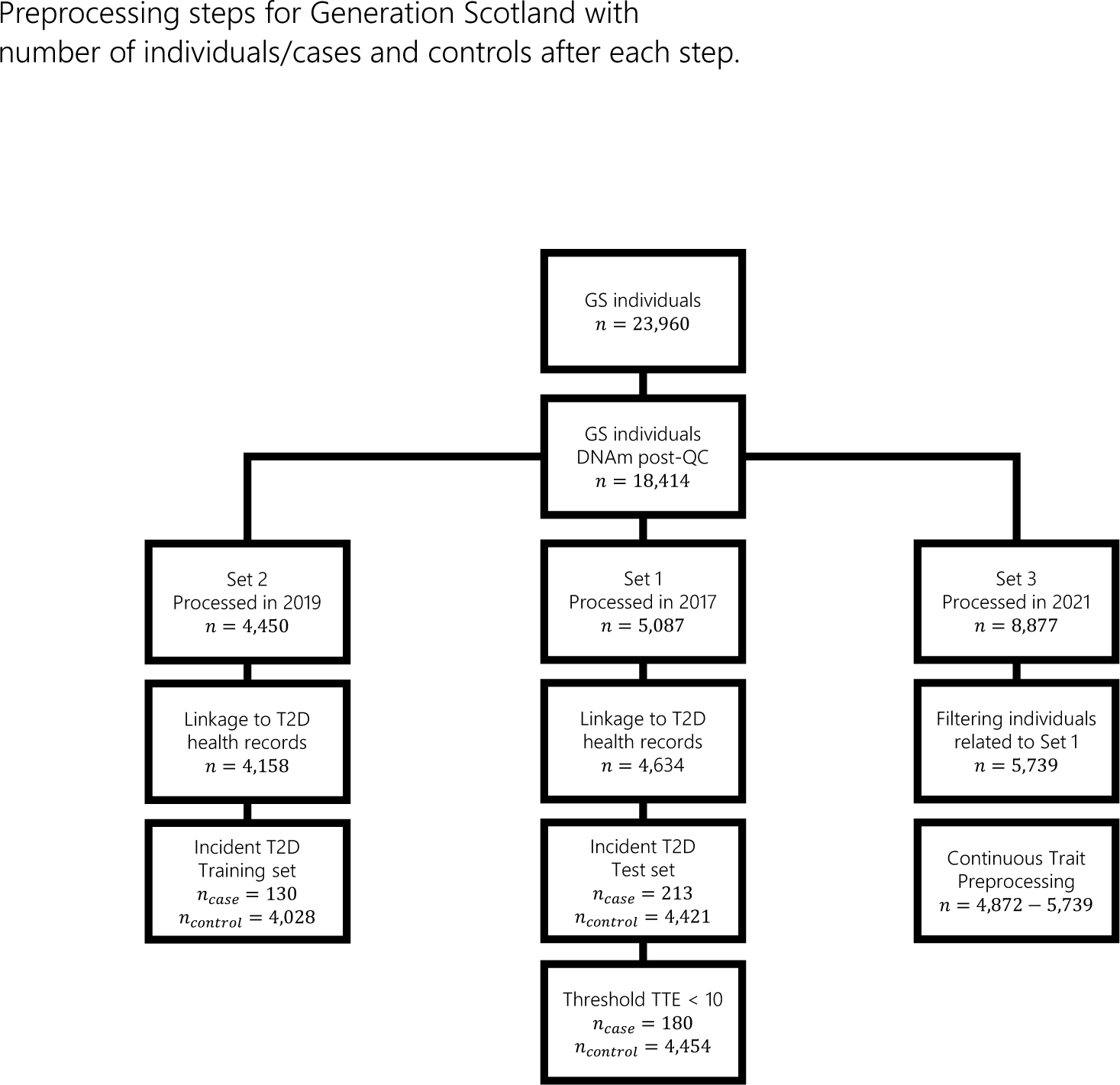
Dataset numbers at each study pipeline processing step

**Supplementary Figure 2:**
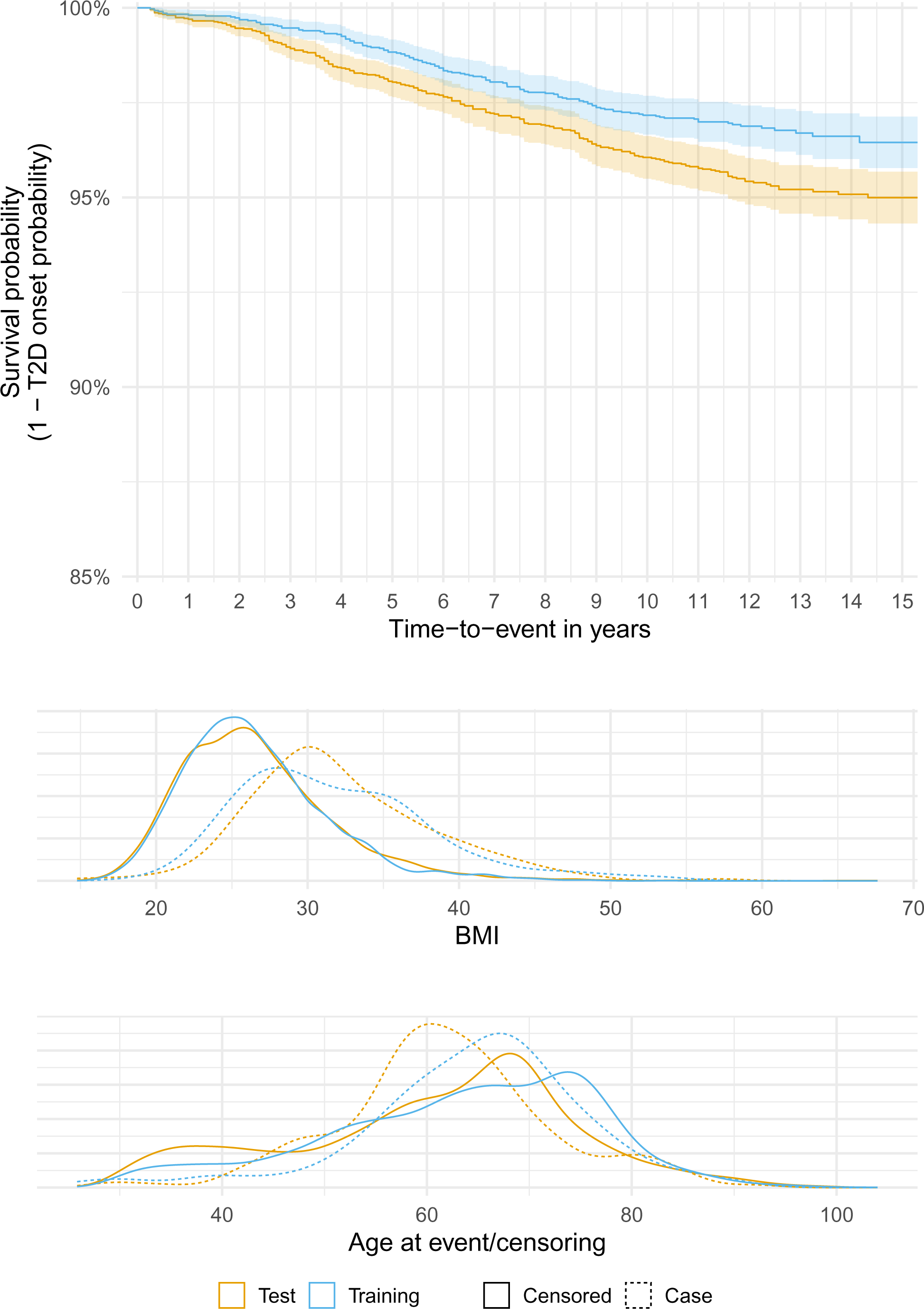
Kaplan-Meier curves (top) and density plots for BMI and age at event/censoring (middle and bottom, respectively) for the incident T2D training and test sets

**Supplementary Figure 3:**
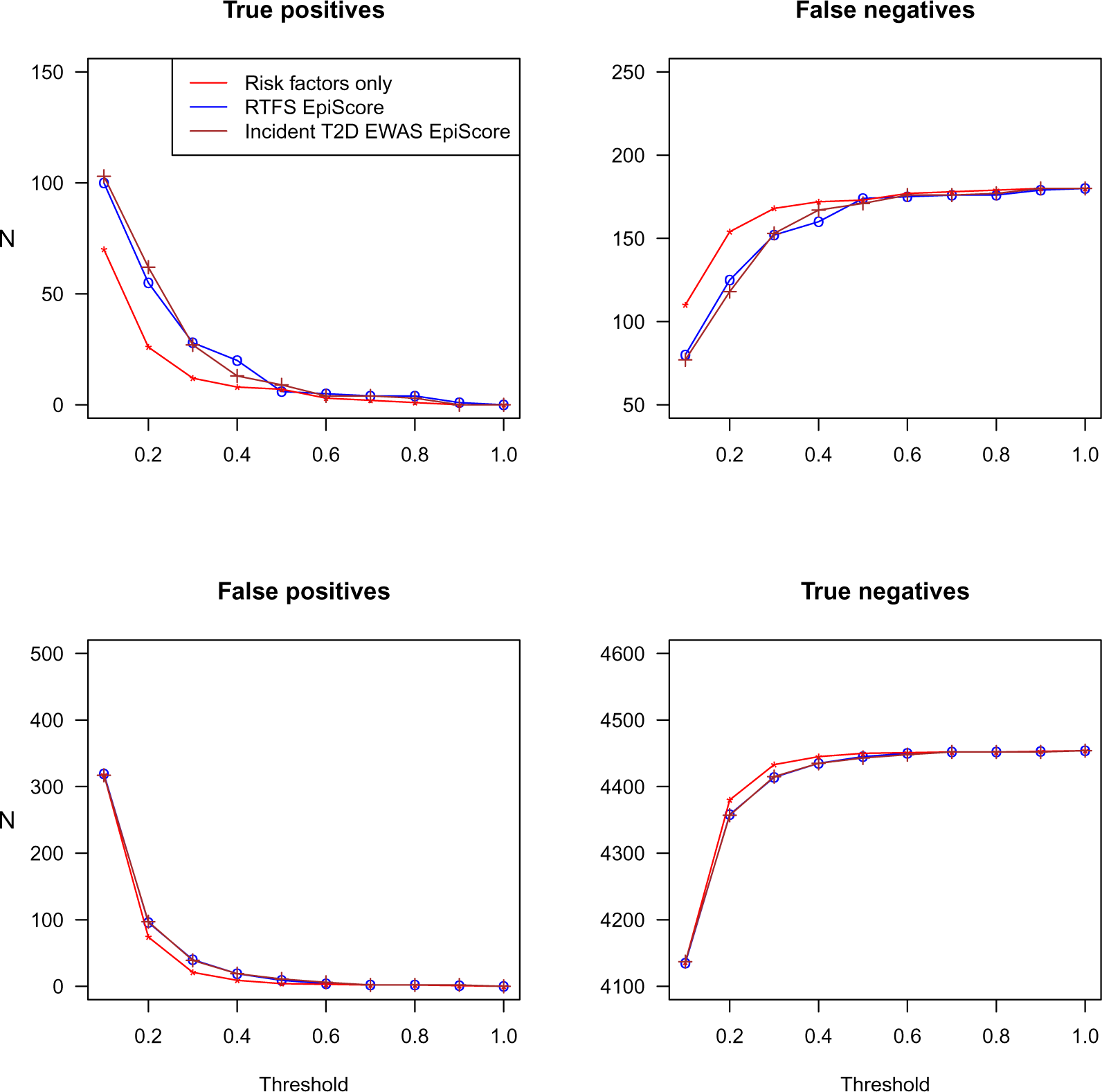
Confusion matrix metrics across the probability threshold range 0-1 for the RTFS, incident T2D EWAS-based filtering and risk factors-only models.

**Supplementary Figure 4:**
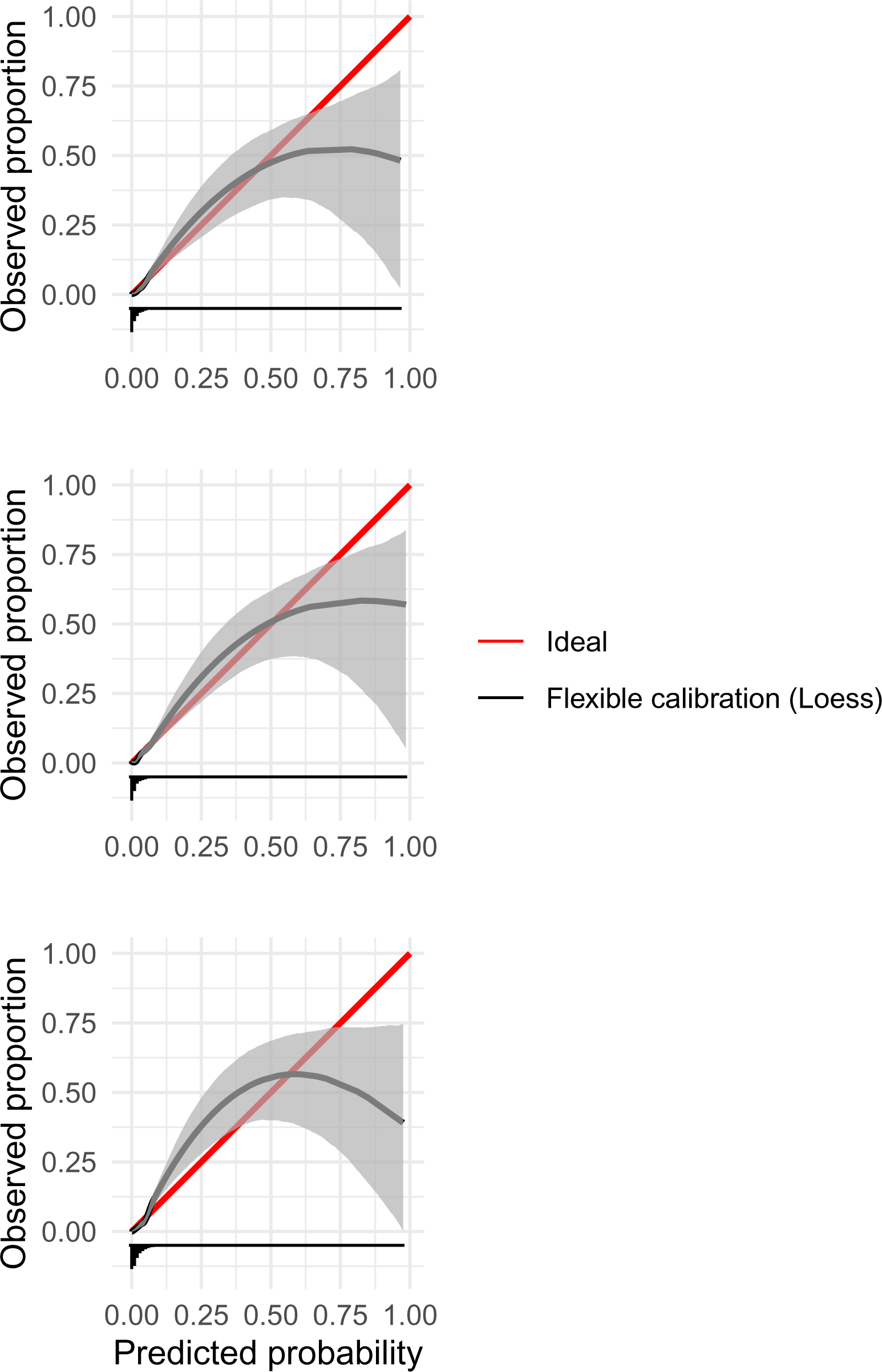
Calibration curves for the incident T2D EWAS EpiScore model (top), RTFS EpiScore model (middle) and risk factors-only model (bottom). The latter shows weaker calibration performance; the fitted calibration curve (black) is overall further from the perfect calibration line (red). All models show underestimation of risk below a predicted probability of around 0.5 and overestimation of risk otherwise.

**Supplementary Figure 5:**
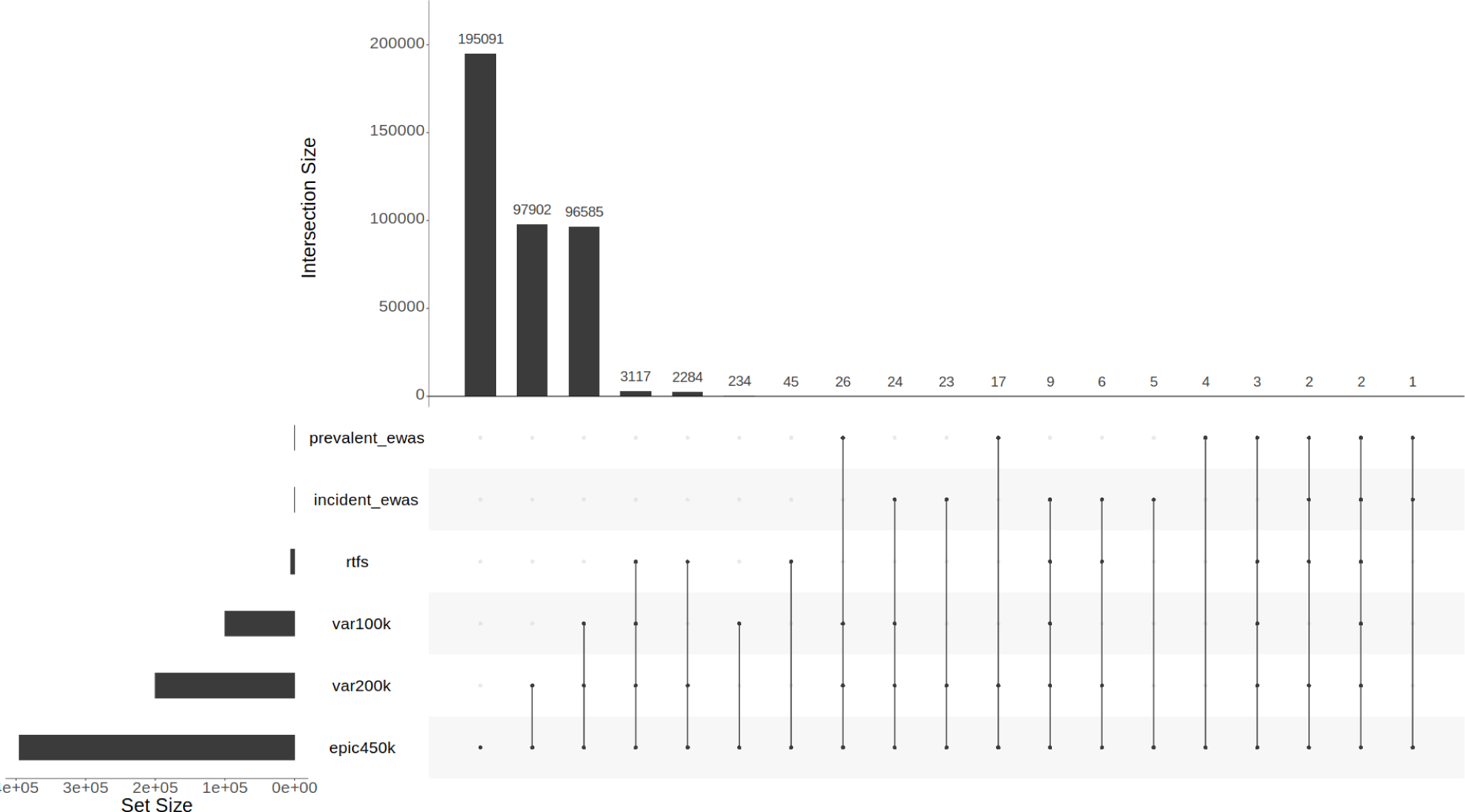
UpSet plot showing number of CpGs selected using each pre-selection method and overlaps between methods. Each column represents the number of CpGs pre-selected by the corresponding specific combination of methods. This was generated with the ”distinct” option, meaning the presence or absence of a point in a column explicitly corresponds to the presence or absence of the corresponding method in the set.

**Supplementary Figure 6:**
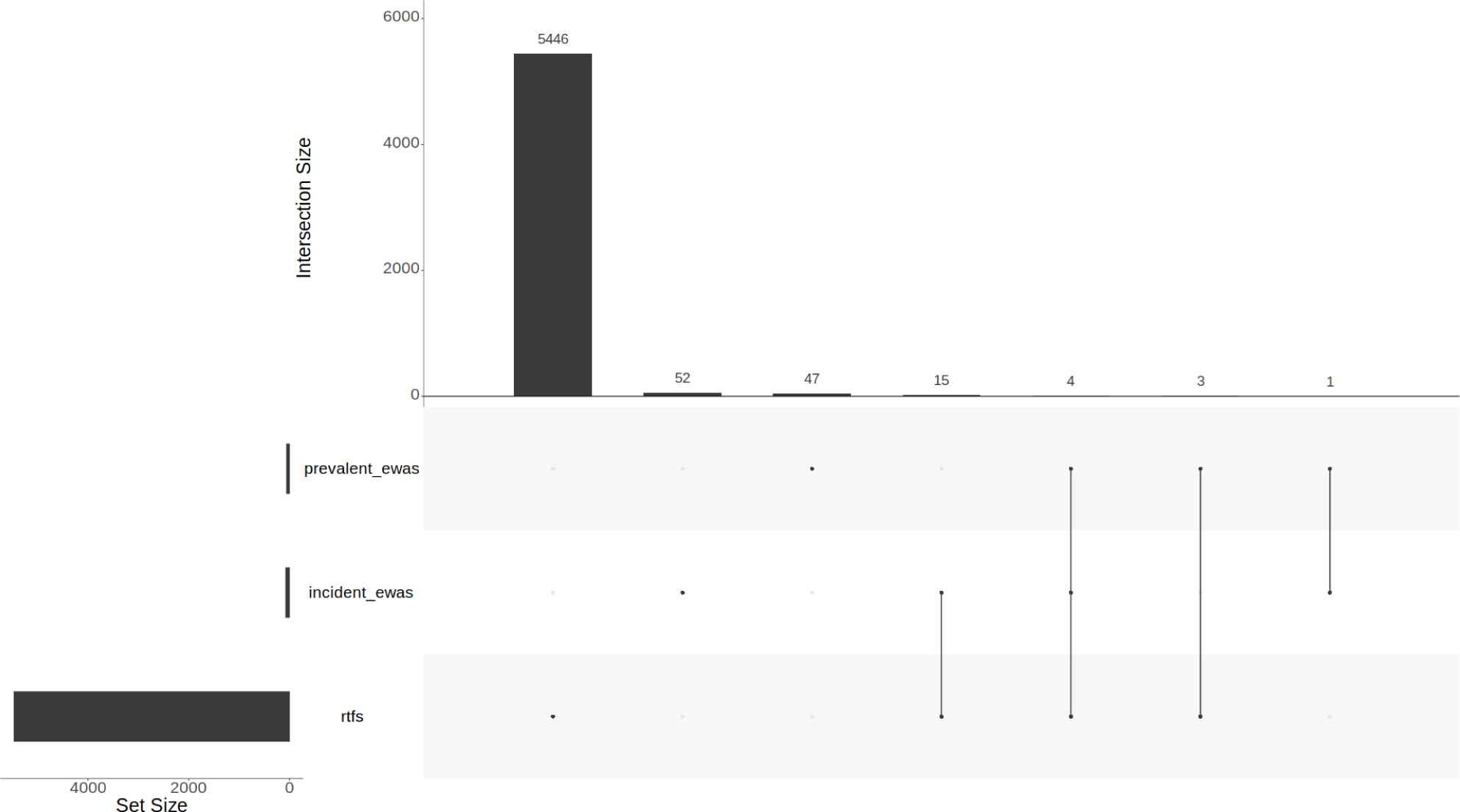
UpSet plot showing number of CpGs selected using the RTFS and EWAS-based pre-selection methods and overlaps between methods. Each column represents the number of CpGs pre-selected by the corresponding specific combination of methods. This was generated with the ”distinct” option, meaning the presence or absence of a point in a column explicitly corresponds to the presence or absence of the corresponding method in the set.

