## Supplementary material for "Feature pre-selection for the development of epigenetic biomarkers": supplementary_note.docx

**Calculation of incremental Cox proportional-hazards (Cox PH) predictions**

Predictions from the incremental Cox PH models can be calculated as follows.

First, calculate the linear predictor:

Null (risk factors-only) model:

$$linear\_predictor_{null}=\beta_{Age}Age+\beta_{Sex}Sex+\beta_{BMI}BMI+\beta_{Hypertension}Hypertension+\beta_{family\_diabetes}family\_diabetes+\beta_{PRS}PRS$$

where each $\beta$ is the corresponding coefficient in the Cox PH model.

Models including an EpiScore:

$$linear\_predictor_{Epi}=linear\_predictor_{null}+\beta_{Epi}Epi$$

where $\beta_{Epi}$ is the corresponding coefficient in the Cox PH model.

$Epi$ is the EpiScore (linear predictor from the Cox PH elastic-net model trained on CpGs) which can be calculated as:

$$Epi=\beta_{CpG1}CpG1+\beta_{CpG2}CpG2+\beta_{CpG3}CpG3+\ldots\beta_{CpGp}CpGp$$

where each $\beta_{CpGx}$ is the corresponding coefficient in the Cox PH elastic-net model.

The cumulative baseline hazard is calculated using the basehaz.gbm function in the gbm R package version 2.1.8. The 10-year survival probability is then calculated as ${S\left( t \right)=exp\left[ -(\Lambda_{0}(t)) \right]}^{exp\left( linear\_predictor \right)}$ at $t=10$, where $\Lambda_{0}(t)$ is the cumulative baseline hazard. The 10-year onset probability is therefore $1-S(10)$.

**Validation of RTFS model in KORA S4**

**KORA S4 cohort information**

The present analyses are based on a subsample of the participants of the KORA S4 study. KORA

(Cooperative Health Research in the Region of Augsburg) is a research platform performing

population-based surveys and subsequent follow-ups in the region of Augsburg in Southern

Germany. Participants were of German nationality, aged between 25-74 years, 50% female,

and sampled from the population registers in the study area where main place of residence was: Ausburg city town, county Ausburg or county Aichach-Friedberg. Each participant completed a health questionnaire, providing details on health status and medication. Blood samples were also taken for assaying of omics data. KORA S4 participants were recruited between 25/10/1999-28/04/2001. This study used a subsample of the 1,451 participants of the KORA S4 study with DNAm and incident T2D data available and no prevalent diabetes at baseline.

**Participant follow-up and ascertainment of incident T2D diagnoses**

For diabetes morbidity, the data are limited to a follow-up of 10 years - starting from KORA S4 recruitment. For incident T2D all prevalent diabetics as well all other diabetes types except T2D cases are excluded. All incident cases of type 2 diabetes which had been diagnosed within a follow up of 10 years were included. Self-reported incident cases of diabetes were validated by hospital records or by contacting the treating physician. Furthermore, the hospital records of those deceased during the follow-up period without a diagnosis of diabetes at baseline were also examined and/or their last treating physician was contacted. The records were searched, or the physicians were asked for a history of diabetes and if a person had suffered from diabetes the type of diabetes and the date of diagnosis were ascertained. Age, BMI, hypertension, sex as well as self-reported family (mother or father)

history of T2D were taken at the baseline of KORA S4. BMI was calculated as the individual's weight in kg divided by the square of their height in metres.
